# Trends, District-Level Variations, and Socioeconomic Disparities in Cesarean Section Delivery and its Association with Neonatal Mortality in Bangladesh

**DOI:** 10.1101/2024.02.26.24303360

**Authors:** Md Nuruzzaman Khan, Md Badsha Alam, Shimlin Jahan Khanam, M Mofizul Islam, Md Arif Billah

**Affiliations:** Department of Population Science, Jatiya Kabi Kazi Nazrul Islam University, Mymensingh, Bangladesh; Department of Public Health, La Trobe University, Melbourne 3086, Australia; Health System and Population Studies Division, International Centre for Diarrheal Disease Research, Bangladesh (icddr,b), 68 Shaheed Tajuddin Ahmed Sarani, Mohakhali, Dhaka 1212, Bangladesh

**Keywords:** Cesarean delivery, Institutional delivery, Neonatal mortality, Bangladesh Demographic and Health Survey, Bangladesh

## Abstract

**Background:** Cesarean section (CS) rates have risen dramatically worldwide, with a majority of the countries exceeding the World Health Organization’s (WHO) preferred rate of 10-15%. However, disparities exist, with evidence suggesting that socioeconomic disadvantage and geographic location play significant roles. Despite this, comprehensive estimates, especially in Bangladesh, remain scarce. This study aims to determine trends, district-level variations, and socioeconomic disparities in CS rates in Bangladesh.

**Methods:** Data from six rounds of Bangladesh Demographic and Health Surveys were analyzed. The considered outcome variables were the occurrence of CS delivery in relation to the mode of delivery and delivery place. Neonatal mortality was also assessed as another outcome variable. Explanatory variables included districts, wealth quintiles, and socio-demographic characteristics. Descriptive statistics were used to provide an over-the-year trend and variation in CS delivery in Bangladesh. Multilevel mixed-effects binary logistic regressions were used to explore predictors of CS delivery and the association between CS and neonatal mortality.

**Results:** Between 1999/2000 and 2017/18, hospital births in Bangladesh increased by 42%, primarily driven by a substantial rise in CS delivery, from 30% to 66%. Private healthcare facilities played a significant role, contributing 80% of the country’s total CS delivery in 2017/18, a substantial increase from 45.5% in 1999/2000. In contrast, CS delivery rates in government healthcare facilities decreased from 49.7% to 15.5% during the same period. Deficient use of CS was reported by women in border and hilly districts, as well as those in the poorest wealth quintile. A clear link between a CS delivery and neonatal mortality was not found.

**Conclusion:** The uneven distribution of CS delivery across districts and socioeconomic groups underscores the need for a more nuanced approach to childbirth. While government efforts to curb unnecessary use of CS have fallen short, this study suggests a one-size-fits-all strategy could worsen disparities. Instead, the focus should shift from mere accessibility to ensuring justified and appropriate utilization, with public healthcare facilities playing a key role in offering safe alternatives.

## Introduction

The prevalence of delivery in cesarean section (CS), a life-saving medical procedure, has globally surged, escalating from approximately 7% in 1990 to a staggering 21% in 2023 [1, 2]. Projections indicate that this rate will increase to 30% by 2030, with the majority of such an increase occurring in low- and lower-middle-income countries (LMICs) [2]. Each of these estimates, current and future, exceeds the recommended 10-15% delivery in CS, with over 10% conferring no discernible advantages to maternal and child health[2–4]. While this rise is attributed to improved surgical safety, evolving medical practices, and the enhancement of socio-economic status, it has triggered discussions about its broader public health implications [5].

On the one hand, CS undeniably enhances maternal and neonatal outcomes in specific scenarios, reducing perinatal mortality and morbidity associated with fetal distress, dystocia, and pre-existing maternal health conditions [6, 7]. However, the overuse of CS exposes both mothers and infants to unnecessary surgical risks, including infections, hemorrhage, and enduring complications [8, 9]. Moreover, it has the potential to disturb the delicate establishment of the newborns’ microbiome, influencing future health [10].

Furthermore, this increasing trend of CS use often prompts a focus on overuse, overlooking the serious lack of access to CS in many LMICs, such as Somalia, or in areas underserved within countries, highlighting the issue of necessity rather than overuse [11, 12]. Additionally, disparities in access to these procedures underscore socio-economic inequalities. Wealthier countries and individuals often exhibit higher rates of use, not necessarily due to a higher incidence of pregnancy complications and other cases where CS is recommended, but rather reflecting disparate needs based on economic status.[13–15] This complexity often creates a dilemma, with the genuine need hidden by overuse, contributing to the rise in maternal and child mortality [15]. Addressing this challenge necessitates segregated data for each country and areas within countries, accompanied by comprehensive lists of factors associated with corresponding CS and how such estimates relate to maternal and child health. However, this is often lacking in LMICs, where data frequently focuses on country-level estimates and socio-demographic factors [3, 15–18].

Bangladesh, an LMIC, records over 4 million annual births, with approximately half delivered through CS, significantly contributing to the global increase in CS [15]. This higher rate of CS is often unjustified, as disadvantaged women—particularly those in rural areas with lower education and socio-economic status—report significantly lower CS utilization, even if they are in need of such service [15]. Conversely, CS is highly concentrated among women with improved socio-economic status and those residing in urban areas, as well as their combinations [3, 15–18]. This indicates the possibility of significant regional variations in CS delivery, supported by existing evidence of differing coverage in terms of rurality, education, and socio-economic status [2, 15]. However, this nuanced understanding is frequently overlooked in country-level policies and programs in LMICs, particularly in Bangladesh, which tend to focus on controlling the overuse of CS nationwide through uniform rules and regulations rather than considering regional estimates [15]. This approach is primarily based on available data from published research, where the lowest geographical unit is a region (the first and largest administrative unit of Bangladesh) that comprises several districts (second administrative unit of Bangladesh) with significant variations within a region in terms of rurality, education, and socio-economic status [3, 15–18]. A uniform policy that does not account for the variations may overlook the field-level needs, make the programs less effective, and create a pathway to increase maternal and child mortality [15]. Our study aims to estimate trends, district-level variations, and socio-economic disparities in CS delivery in Bangladesh. The study also examined the association between cesarean rates and neonatal mortality rates. Previous studies found a negative association between these two[19–21]; we want to see if the association holds in the context of Bangladesh.

## Methods

### Study design and sampling technique

We conducted a comprehensive analysis using data from six rounds of the Bangladesh Demographic and Health Survey (BDHS) spanning the years 1999/2000 to 2017/18, each conducted at three-to-four-year intervals. BDHS is a nationally representative household survey, an integral component of the Demographic and Health Survey Program administered by the USA in 89 LMICs. Financial support for these surveys was provided by the United States Agency for International Development (USAID) and technical assistance was rendered by ICF International in Calverton, USA. The survey sample comprised of households selected through a two-stage stratified random sampling technique. In the initial stage, primary sampling units (PSUs) were chosen across the country, utilizing PSU lists derived from the most recent national population census conducted by the Bangladesh Bureau of Statistics. Subsequently, a household listing operation was executed, and 30 households were randomly selected from each PSU in the second stage of sampling. Data collection involved surveying women who met specific inclusion criteria: (i) married women of reproductive age (15-49 years old), and (ii) those who were either usual residents of the selected households or had spent the most recent night at the selected households on the day of the survey. Additionally, data were collected for their partners and children under the age of five. For further details, a comprehensive overview of these surveys can be found in the respective survey reports [22–27].

### Analytical sample

We analyzed data of a total of 35,125 participants derived from the six rounds of the BDHS. Of these participants, 6,939 were from the 1999/2000 BDHS, 7,002 from the 2004 BDHS, 6,058 from the 2007 BDHS, 4,956 from the 2011 BDHS, 4,904 from the 2014 BDHS, and 5,266 from the 2017/18 BDHS. The inclusion criteria for sample selection were as follows: (i) having given birth to at least one child within three years of the survey, and (ii) providing information on delivery methods and the place of delivery.

### Outcome variable

The outcome variable was childbirth through a CS, categorized dichotomously as either "yes" or "no". Relevant data were obtained by posing the question to eligible respondents: "*Was the (name) child delivered by cesarean section, that is, did they cut your belly open to take the baby out?*" The response options were yes or no, and these were the parameters considered in this study. We analyzed these responses to estimate two measures: (i) population-level cesarean rates and (ii) institutional cesarean rates. In both cases, the numerator was the occurrence of cesarean section delivery. However, the denominator considered for population-level cesarean rates was the total number of births, including both home and healthcare facilities, while for institutional cesarean rates, it was the total number of births occurring at healthcare facilities. Institutional cesarean births were further classified by the type of facility, such as public, private, and NGO facilities.

### Explanatory variables

The survey data included information on participants under administrative divisions. However, it also included the geographical location points from where the data were collected and provided a unique opportunity to extract district-level data. We linked these locations into a district-level shape file, creating the district variable. The wealth quintile was another explanatory variable and was created based on household assets, such as roof types and ownership of televisions, using principal component analysis. The scores generated through this method were classified into five equal groups with cutoff values at every 0.20, designated as poorest, poorer, middle, richer, and richest. Other explanatory variables included respondents’ socio-demographic characteristics, namely women’s age at birth, women’s education, women’s employment status, the sex of the child, exposure to mass media, place of residence, and regional location. We also included neonatal mortality, defined as the mortality of a child within one month of birth, as an additional explanatory variable.

### Statistical analysis

We initially estimated the rates of CS delivery at the population level, institutional level, government healthcare facility level, private healthcare facility level, and NGO healthcare facility level. CS rates were also examined across districts and wealth quintiles. Moreover, we measured the risk ratio of CS delivery by comparing the risk among the women in the lowest and richest wealth quintiles. Excess or deficit use of CS delivery for each of the districts was identified by comparing the prevalence of CS delivery for the most recent survey (2017/18 BDHS) with the ideal rate of 10-15%, as considered by the international healthcare community[28]. Therefore, districts that had a higher prevalence of CS than 15% were identified as “excess”, within the range of 10-15% were identified as “within range”, and less than 10% were identified as “deficit” categories. We also estimated the district-wise neonatal mortality (deaths within 28 days if livebirths) and compared with the CS rate of the corresponding districts. We employed multilevel mixed-effect binary logistic regressions to investigate factors associated institutional delivery, CS delivery at the population level, and CS delivery at the institutional level. Additionally, the association between CS and neonatal mortality, adjusted for socio-demographic factors, was determined using the same regression approach. The rationale for utilizing multilevel regressions is the hierarchical structure of the BDHSs, where individuals are nested within households, and households are nested within clusters. We adopted a three-level multilevel modeling approach: households, clusters, and survey years. All results were weighted by sampling weights to correct for sample design and are presented with a 95% confidence interval. The entire analysis was conducted using STATA software, while Microsoft Excel was used for constructing graphs, and ArcGIS 10.5 for producing maps. This study adhered to the Strengthening the Reporting of Observational Studies in Epidemiology (STROBE) reporting guidelines.

### Ethical consideration

The data analyzed in this study were obtained from the Demographic and Health Survey Program of the USA. Prior to conducting the survey in Bangladesh, approval was obtained from the institutional review board of the ICF, USA, and subsequently from the National Research Ethics Committee of the Bangladesh Medical Research Council. Informed written consent was obtained from all individuals involved. We obtained permission to access the data for analytical purposes, and the survey authority provided us with deidentified data. As the study involved secondary data analysis and adhered to the relevant guidelines and regulations, no additional ethical approval was required.

## Results

### Background characteristics of the respondents

A majority of the women are within the age range of 20-34 years and have attained either primary or secondary level education. Around two-third of the total women reported that they were not engaged in any formal income-generating activities. Women living in rural areas constitute 73-84% of the sample. The detailed background characteristics of the respondents are presented in Supplementary Table 1.

### Trends in institutional and cesarean delivery rates from 1990/2000 to 2017/18

The trends in institutional and CS deliveries in Bangladesh from 1999/2000 to 2017/18 are presented in Table 1. Institutional deliveries increased by almost 42% (from 8% to 49.8%) over the specified time frame. Concurrently, population-level CS deliveries demonstrated a growth of approximately 30%, escalating from 2.4% to 32.9%. The predominant factor contributing to the overall increase in CS was institutional deliveries. Nearly 66% of institutional deliveries took place in CS, marking a significant rise from the 30% recorded in 1999/2000. There was a substantial surge in CS delivery in private healthcare facilities. In 2017/18, approximately 80% of total births in private healthcare facilities were delivered via CS, representing a twofold increase from the 1999/2000 CS rate in private healthcare facilities (45.5%). The CS rate in healthcare facilities operated by NGOs remained mostly unchanged over the years, while the CS rate in government healthcare facilities declined more than threefold, decreasing from 49.7% in 1999/2000 to 15.5% in 2017/18.

**Table 1.** Trends in institutional delivery and cesarean delivery rates in Bangladesh, 1999/2000 to 2017/18.

| Survey rounds | Delivery rate, % (95% CI) |  | Institutional cesarean, % (95% CI) |  |  |  |
| --- | --- | --- | --- | --- | --- | --- |
|  | Institutional | Population-level cesarean | Total | Government facilities | Private health facilities | NGO health facilities |
| BDHS 1999/2000 | 8.0 (6.9-9.2) | 2.4 (2.0-2.9) | 30.0 (26.0-34.4) | 49.7 (42.2-57.2) | 45.5 (38.0-53.3) | 4.8 (2.3-9.5) |
| BDHS 2004 | 9.9 (8.7-11.2) | 3.5 (2.9-4.1) | 35.1 (30.9-39.5) | 49.0 (41.1-56.9) | 47.2 (40.1-54.5) | 3.8 (1.3-10.5) |
| BDHS 2007 | 14.6 (13.1-16.4) | 7.5 (6.5-8.7) | 51.5 (47.0-56.0) | 32.4 (27.4-37.9) | 62.1 (56.4-67.4) | 5.5 (3.5-8.4) |
| BDHS 2011 | 28.8 (26.8-31.0) | 17.1 (15.6-18.7) | 59.2 (56.0-62.2) | 29.4 (25.4-33.8) | 67.2 (62.9-71.3) | 3.4 (2.2-5.2) |
| BDHS 2014 | 37.6 (34.8-40.5) | 22.9 (20.9-25.0) | 60.9 (58.0-63.6) | 20.9 (17.6-24.6) | 76.3 (72.4-79.9) | 2.8 (1.8-4.4) |
| BDHS 2017/18 | 49.8 (47.5-52.2) | 32.9 (30.9-34.9) | 66.0 (63.7-68.2) | 15.5 (13.5-17.7) | 79.8 (77.4-82.0) | 4.7 (3.7-6.0) |

### Variations in institutional and cesarean section delivery rates across districts

The district-level variations in institutional and CS delivery rates in 2017/18 survey are presented in Table 2. The last column indicates whether the CS rates are “excess”, “deficit” or “within range” when compared with the population level recommended utilization rate of 15%. Meherpur district recorded the highest rate of institutional delivery (95.0%, 95% CI: 81.8–98.8), while Khagrachari district recorded the lowest rate (10.5%, 95% CI: 1.4–49.6). Generally, districts in the central region of Bangladesh had higher institutional delivery rates, with those near national borders showing lower rates. Moreover, hilly areas, namely Khagrachari (17.7%), Bandarban (10.5%), and Rangamati (22.8%), displayed lower institutional delivery rates.

**Table 2.** Institutional delivery and cesarean section delivery rates by districts, Bangladesh, 2017/18.

| Districts | Delivery rate, % (95% CI) |  | Institutional cesarean, % (95% CI) |  |  |  |  |
| --- | --- | --- | --- | --- | --- | --- | --- |
|  | Institutional | Population-level caesarean | Total | Government facilities | Private health facilities | NGO health facilities | Deficit or excess in comparison with WHO's recommendation of 10-15% use of CS |
| Bagerhat | 47.9 (27.0-69.6) | 23.0 (11.4-40.9) | 48.0 (31.1-65.4) | 26.6 (9.8-54.9) | 73.4 (45.2-90.2) | 0 | Excess |
| Bandarban | 17.7 (11.7-23.3) | 0 | 0 | 0 | 0 | 0 | Deficit |
| Barguna | 42.8 (31.0-55.5) | 34.2 (24.0-46.2) | 79.9 (64.7-89.7) | 12.1 (4.7-27.7) | 87.9 (72.3-95.3) | 0 | Excess |
| Barishal | 47.3 (35.1-59.8) | 36.0 (26.9-46.3) | 76.1 (66.6-83.6) | 20.0 (9.2-38.2) | 76.0 (59.8-87.1) | 4.0 (1.0-14.8) | Excess |
| Bhola | 27.3 (19.9-36.3) | 12.2 (7.2-20.0) | 44.7 (27.5-63.3) | 21.7 (6.9-51.0) | 71.7 (43.1-89.3) | 6.8 (0.9-38.1) | In range |
| Bogura | 45.8 (32.6-59.7) | 30.3 (23.3-38.3) | 66.0 (53.0-77.0) | 27.0 (12.3-49.4) | 68.3 (47.4-83.7) | 4.7 (1.4-14.6) | Excess |
| Brahamanbaria | 48.1 (38.0-58.3) | 29.4 (23.9-35.6) | 61.1 (51.9-69.5) | 7.2 (1.0-36.4) | 92.8 (63.6-99.0) | 0 | Excess |
| Chandpur | 52.5 (47.1-57.9) | 40.0 (34.0-46.3) | 76.2 (58.3-88.0) | 6.7 (1.6-24.0) | 93.3 (76.0-98.4) | 0 | Excess |
| Chittagong | 58.7 (44.4-71.7) | 26.2 (17.3-37.7) | 44.7 (33.8-56.1) | 24.3 (13.4-39.9) | 72.4 (57.8-83.4) | 3.3 (0.8-12.8) | Excess |
| Chuadanga | 73.0 (59.4-83.4) | 54.0 (45.2-62.6) | 74.0 (55.5-86.6) | 11.6 (3.4-32.4) | 88.4 (67.6-96.6) | 0 | Excess |
| Cumilla | 50.7 (40.0-61.4) | 38.8 (29.6-48.8) | 76.5 (61.5-86.9) | 10.7 (5.2-20.8) | 86.7 (75.7-93.1) | 2.6 (0.4-17.0) | Excess |
| Cox's Bazar | 38.3 (25.2-53.4) | 15.7 (8.3-27.8) | 41.1 (20.5-65.3) | 8.4 (1.7-32.4) | 83.1 (44.5-96.8) | 8.4 (1.7-32.4) | Excess |
| Dhaka | 72.6 (66.9-77.7) | 50.3 (44.1-56.6) | 69.3 (63.2-74.8) | 24.4 (17.2-33.2) | 68.4 (58.6-76.7) | 7.2 (3.9-12.9) | Excess |
| Dinajpur | 67.2 (49.0-81.4) | 44.8 (33.3-56.9) | 66.7 (56.9-75.3) | 6.5 (2.0-19.0) | 85.4 (63.1-95.2) | 8.1 (2.2-25.9) | Excess |
| Faridpur | 51.6 (24.1-78.1) | 39.2 (21.7-60.0) | 76.1 (65.0-84.5) | 19.6 (9.3-36.7) | 80.4 (63.3-90.7) | 0 | Excess |
| Feni | 57.0 (39.9-72.6) | 34.1 (24.5-45.2) | 59.8 (48.3-70.3) | 16.3 (5.9-37.6) | 77.4 (68.1-84.6) | 6.4 (1.5-23.3) | Excess |
| Gaibandha | 46.0 (33.2-59.2) | 17.2 (8.1-32.9) | 37.4 (20.1-58.7) | 9.9 (2.8-29.5) | 90.1 (70.6-97.2) | 0 | Excess |
| Gazipur | 54.6 (43.7-65.1) | 33.9 (24.1-45.3) | 62.1 (41.7-79.0) | 22.0 (11.6-37.8) | 72.6 (59.9-82.5) | 5.3 (0.8-27.5) | Excess |
| Gopalganj | 42.7 (12.9-78.9) | 36.6 (11.8-71.3) | 85.7 (85.7-85.7) | 16.7 (16.7-16.7) | 83.3 (83.3-83.3) | 0 | Excess |
| Habiganj | 28.6 (16.3-45.2) | 15.5 (7.7-28.7) | 54.0 (36.4-70.6) | 37.1 (18.3-60.8) | 54.2 (29.8-76.7) | 8.7 (2.2-29.2) | Excess |
| Jamalpur | 38.5 (28.7-49.3) | 27.8 (18.4-39.8) | 72.4 (54.1-85.3) | 13.3 (5.0-30.8) | 86.7 (69.2-95.0) | 0 | Excess |
| Jessore | 70.3 (58.1-80.2) | 51.3 (39.0-63.4) | 72.9 (54.7-85.7) | 19.0 (5.8-47.1) | 73.0 (48.5-88.6) | 8.0 (2.4-23.1) | Excess |
| Jhalokati | 51.4 (41.0-61.6) | 31.4 (24.4-39.8) | 61.1 (37.8-80.2) | 10.9 (1.8-45.5) | 89.1 (54.5-98.2) | 0 | Excess |
| Jhenaidah | 64.6 (46.5-79.3) | 45.8 (24.4-82.0) | 70.9 (58.9-80.6) | 0 | 100 | 0 | Excess |
| Joypurhat | 79.7 (65.2-89.2) | 47.6 (27.9-68.1) | 59.7 (26.3-86.1) | 4.9 (0.6-31.2) | 95.1 (68.8-99.4) | 0 | Excess |
| Khagrachhari | 10.5 (1.4-49.6) | 0 | 0 | 0 | 0 | 0 | Deficit |
| Khulna | 60.1 (45.0-73.4) | 43.8 (29.2-59.5) | 72.9 (58.1-83.9) | 12.2 (4.2-30.7) | 75.3 (61.6-85.3) | 12.4 (6.4-22.7) | Excess |
| Kishoreganj | 41.3 (26.3-58.0) | 32.8 (23.2-44.0) | 79.5 (65.1-88.9) | 21.9 (10.1-41.3) | 65.7 (52.9-76.6) | 12.4 (4.1-31.6) | Excess |
| Kurigram | 30.7 (17.5-48.1) | 19.8 (11.6-31.9) | 64.7 (50.2-76.9) | 10.9 (3.7-30.6) | 83.8 (58.4-95.0) | 5.4 (0.9-25.1) | Excess |
| Kushtia | 57.7 (48.5-66.4) | 45.2 (35.7-55.2) | 78.5 (64.0-88.2) | 0 | 87.2 (70.1-95.2) | 12.8 (4.8-29.9) | Excess |
| Lakshmipur | 31.8 (12.8-59.7) | 20.1 (6.7-46.6) | 63.1 (30.6-87.0) | 0 | 100 | 0 | Excess |
| Lalmonirhat | 32.4 (12.7-61.2) | 18.8 (9.7-33.2) | 58.0 (29.3-82.1) | 0 | 100 | 0 | Excess |
| Madaripur | 45.6 (35.1-58.7) | 41.7 (26.7-58.3) | 91.4 (74.6-97.4) | 27.3 (12.4-50.0) | 72.7 (50.0-87.6) | 0 | Excess |
| Magura | 46.1 (35.7-56.8) | 28.7 (18.9-41.1) | 62.4 (41.5-79.5) | 28.3 (13.3-39.5) | 71.7 (60.5-80.7) | 0 | Excess |
| Manikganj | 37.6 (9.5-77.6) | 26.9 (6.7-65.4) | 71.6 (42.9-89.4) | 0 | 100 | 0 | Excess |
| Maulvibazar | 36.0 (24.7-49.2) | 17.3 (10.8-26.4) | 47.9 (36.3-59.8) | 17.3 (5.9-41.2) | 76.9 (52.9-90.8) | 5.8 (0.8-33.2) | Excess |
| Meherpur | 95.0 (81.8-98.8) | 67.5 (63.5-71.2) | 71.0 (65.9-75.7) | 8.3 (1.7-31.3) | 84.3 (77.8-89.1) | 7.4 (1.7-26.7) | Excess |
| Munshiganj | 74.9 (57.1-87.0) | 61.6 (47.1-74.3) | 82.2 (59.0-93.7) | 8.8 (3.6-17.3) | 91.2 (80.4-96.4) | 0 | Excess |
| Mymensingh | 47.7 (40.7-54.7) | 32.3 (26.8-38.3) | 67.7 (58.8-75.6) | 9.6 (4.7-18.5) | 83.5 (70.8-91.3) | 7.0 (2.8-16.1) | Excess |
| Naogaon | 63.6 (46.6-77.7) | 50.9 (33.6-68.1) | 80.1 (61.7-90.9) | 12.9 (5.5-27.2) | 87.1 (72.9-94.5) | 0 | Excess |
| Narail | 47.2 (44.0-50.5) | 36.2 (29.7-43.3) | 76.7 (52.6-90.7) | 39.1 (26.8-52.9) | 60.9 (47.1-73.2) | 0 | Excess |
| Narayanganj | 64.9 (54.7-73.9) | 54.8 (47.5-61.9) | 84.5 (55.2-96.0) | 23.5 (8.8-49.4) | 70.8 (51.2-84.8) | 5.7 (1.0-26.7) | Excess |
| Narsingdi | 51.6 (28.0-74.4) | 42.3 (21.9-65.7) | 82.1 (64.1-92.1) | 16.5 (4.8-43.5) | 80.0 (54.4-93.1) | 3.6 (0.4-23.2) | Excess |
| Natore | 63.4 (38.1-83.0) | 41.8 (27.6-57.5) | 66.0 (49.2-79.5) | 6.4 (1.2-27.6) | 93.6 (72.5-98.8) | 0 | Excess |
| Chapai Nawabganj | 44.4 (38.1-51.0) | 23.2 (15.0-34.1) | 52.2 (35.2-68.8) | 0 | 100 | 0 | Excess |
| Netrakona | 29.1 (20.5-39.6) | 19.6 (13.0-28.5) | 67.3 (49.6-81.1) | 5.5 (1.3-20.0) | 94.5 (80.0-98.7) | 0 | Excess |
| Nilphamari | 52.5 (40.6-65.4) | 22.8 (11.6-39.6) | 42.6 (21.7-66.6) | 0 | 86.7 (72.8-94.1) | 13.3 (5.9-27.2) | Excess |
| Noakhali | 41.1 (25.7-58.4) | 21.9 (12.3-36.0) | 53.4 (42.7-63.7) | 18.0 (9.5-31.5) | 82.0 (68.5-90.5) | 0 | Excess |
| Pabna | 46.9 (36.6-57.3) | 27.1 (18.9-37.2) | 57.8 (42.7-71.5) | 7.4 (1.6-28.7) | 92.6 (71.3-98.4) | 0 | Excess |
| Panchagarh | 25.9 (15.3-40.3) | 16.1 (5.8-37.2) | 62.1 (28.3-87.2) | 13.2 (2.8-44.6) | 86.8 (55.4-97.2) | 0 | Excess |
| Patuakhali | 31.7 (18.1-49.2) | 16.5 (8.8-28.7) | 52.1 (38.4-65.5) | 11.8 (3.7-32.0) | 88.2 (68.0-96.3) | 0 | Excess |
| Pirojpur | 51.2 (37.9-64.3) | 30.2 (19.6-43.4) | 59.0 (47.5-69.6) | 9.9 (3.8-23.8) | 90.1 (76.2-96.2) | 0 | Excess |
| Rajbari | 80.8 (43.5-95.8) | 61.9 (26.8-87.8) | 76.6 (53.8-90.2) | 0 | 89.7 (49.8-98.7) | 10.3 (1.3-50.2) | Excess |
| Rajshahi | 80.1 (64.7-89.8) | 59.1 (45.9-71.1) | 73.8 (60.5-83.8) | 15.8 (5.3-38.8) | 77.7 (53.3-91.4) | 6.5 (1.4-25.3) | Excess |
| Rangamati | 22.8 (4.6-64.6) | 11.4 (2.6-38.6) | 50.0 (50.0-50.0) | 33.3 (33.3-33.3) | 66.7 (66.7-66.7) | 0 | In range |
| Rangpur | 43.9 (25.6-64.1) | 31.6 (17.8-49.5) | 71.9 (58.0-82.6) | 6.8 (2.4-17.3) | 73.0 (62.1-81.7) | 20.2 (11.3-33.6) | Excess |
| Satkhira | 60.5 (40.5-77.5) | 42.0 (22.4-64.6) | 69.5 (42.9-87.3) | 10.5 (2.8-32.7) | 87.2 (63.3-96.4) | 2.2 (0.3-14.3) | Excess |
| Shariatpur | 30.7 (11.2-60.8) | 20.4 (4.9-55.8) | 66.3 (36.5-87.1) | 21.3 (2.4-75.0) | 78.7 (25.0-97.6) | 0 | Excess |
| Sherpur | 23.8 (13.5-38.5) | 10.2 (3.6-25.6) | 43.0 (19.6-69.9) | 28.6 (8.8-62.4) | 65.4 (35.1-86.8) | 6.1 (1.1-28.4) | In range |
| Sirajganj | 37.9 (29.1-47.6) | 29.3 (17.7-44.3) | 77.3 (49.4-92.2) | 14.4 (4.2-39.0) | 80.0 (57.3-92.3) | 5.6 (1.5-18.5) | Excess |
| Sunamganj | 31.9 (21.2-44.9) | 17.6 (10.9-27.2) | 55.2 (44.1-65.8) | 47.8 (26.9-69.6) | 42.5 (21.9-66.1) | 9.7 (4.1-21.0) | Excess |
| Sylhet | 52.5 (42.6-62.2) | 35.1 (27.2-43.9) | 66.8 (55.2-76.8) | 24.7 (15.3-37.4) | 64.2 (50.7-75.7) | 11.1 (4.8-23.4) | Excess |
| Tangail | 49.4 (36.4-62.5) | 38.3 (28.4-49.3) | 77.5 (59.2-89.1) | 13.0 (3.9-35.7) | 87.0 (64.3-96.1) | 0 | Excess |
| Thakurgaon | 72.6 (63.4-80.2) | 45.2 (30.5-60.9) | 62.3 (37.9-81.7) | 3.2 (0.4-21.4) | 91.4 (68.4-98.1) | 5.3 (0.7-30.8) | Excess |

**Note:** Presented as row percentage

These trends were mirrored in overall CS rates (Supplementary figure 1), although specific figures varied across healthcare facilities (Supplementary figures 2, 3). Some districts reported no CS occurrences in public healthcare facilities, like Khagrachari, Bandarban, Nilphamari, Chapai-Nawabganj, Lalmonirhat, Laksmipur, Jenaidah, Manikganj, Rajbari, and Kushtia, indicating all CS cases in these districts occurred in private healthcare facilities.

Conversely, public healthcare facilities in certain districts, such as Sunamganj, reported CS delivery rates two to three times higher than the national average rate for public healthcare facilities. Despite variations, private healthcare facilities dominated CS procedures in all districts, accounting for up to 90% of cases.

The rise in CS rates has been substantial in recent years. Comparing findings from the two most recent surveys, BDHS 2014 and BDHS 2017/18, we found that for 54 of the total 64 districts, population-level CS rates increased by up to 40.4% points (in Rajbari), while in 6 districts (Satkhira, Gopalgonj, Makingionj, Maulvibazar, Panchagarh, and Sherpur), rates declined by up to 14.4% points. Bandarban and Khagrachari reported no CS births (Supplementary Table 2). CS rates in NGO-operated healthcare facilities ranged from 0 to 20.2%, with the highest rate reported in Rangpur (20.2%, 95% CI: 11.3–33.6).

Overall, 59 of the total 64 districts in Bangladesh reported excess use, two districts (Bandarban, Khagrachari) reported deficit use, and three districts (Bhola, Rangamati, and Sherpur) reported CS delivery rates within the recommended range (Table 2).

### Socio-economic differentials of cesarean delivery across districts

The rates of CS delivery across wealth quintiles are presented in Table 3. Among the women in poorest quintile, the CS delivery rate was 13.1% (95% CI: 10.7–15.9) and among the women in the richest quintile, it was 61.5% (95% CI: 57.8–64.9). This disparity was noticeable across districts, with consistently higher rates of CS reported by women in the richest quintile. In almost half of the districts (n=32), the proportion of CS deliveries among the poorest quintile was below the 15% threshold, and 30 of them were below the 10% threshold. Some districts reported no CS deliveries, while in Meherpur, all births among the poorest quintile were delivered via CS. Conversely, for the richest quintile, nearly all districts had CS rates exceeding 40%, except Habiganj (35.8%), Pirojpur (38.8%), and Sunamganj (34.6%), while Manikganj and Rangamati reported no CS among the richest clusters, despite the existence of total CS rates. When comparing the poorest and the richest in urban and rural areas, significant variations were evident. There were substantial variations in risk ratios of CS delivery between the poorest and richest quintiles, ranging from <2.0 (Nawabganj, Gopalganj, Rajshahi, Pabna, Jessore, and Brahmanbaria) to 18.5 (Rangpur) (Figure 1).

**Figure 1.**
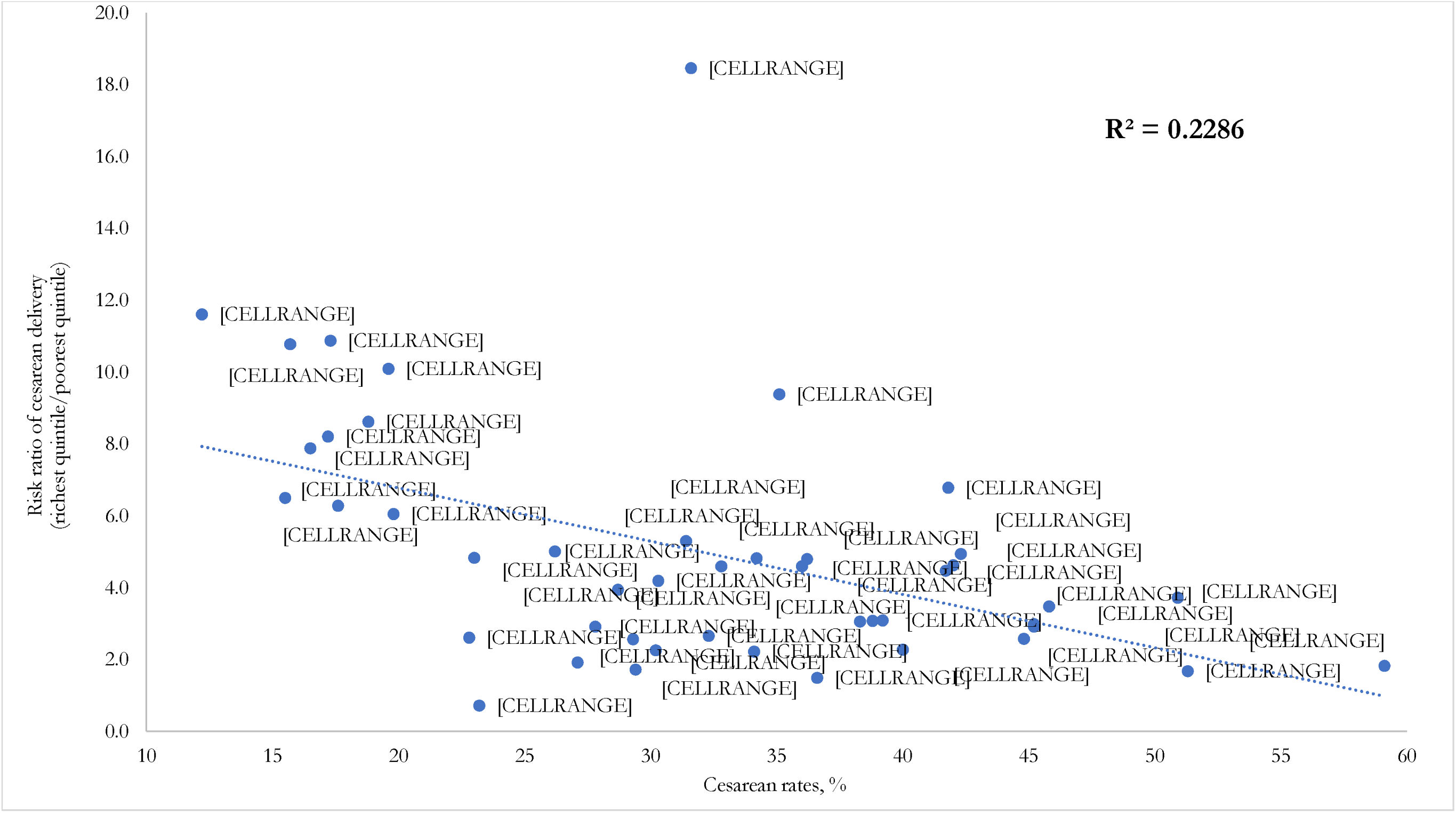
Cesarean rates and risk ratio of cesarean section deliveries between the poorest and richest quintiles across districts (BDHS 2017/18)

**Table 3.** Population-based cesarean section rates by socioeconomic quintile and districts, Bangladesh, 2017/18.

| Districts | Poorest | Poorer | Middle | Richer | Richest |
| --- | --- | --- | --- | --- | --- |
|  | % (95% CI) | % (95% CI) | % (95% CI) | % (95% CI) | % (95% CI) |
| Bagerhat | 10.7 (1.5-48.8) | 27.1 (9.1-58.1) | 40.5 (21.9-62.3) | 10.6 (1.8-43.7) | 51.8 (5.9-94.9) |
| Bandarban | 0 | 0 | 0 | 0 | 0 |
| Barguna | 20.7 (10.2-37.4) | 25.9 (9.4-36.0) | 52.0 (24.0-78.8) | 58.0 (29.5-82.1) | 100 |
| Barishal | 18.6 (6.8-41.7) | 18.9 (8.8-36.0) | 28.8 (12.6-53.2) | 60.2 (40.3-77.2) | 85.7 (91.5-95.7) |
| Bhola | 6.0 (1.7-19.2) | 5.9 (1.6-19.1) | 25.1 (15.9-37.2) | 8.3 (1.0-43.7) | 69.7 (30.3-92.4) |
| Bogura | 10.4 (3.0-30.4) | 27.7 (14.4-46.5) | 41.8 (23.2-63.0) | 36.2 (19.7-56.8) | 43.7 (20.5-70.1) |
| Brahmanbaria | 24.7 (8.4-53.8) | 6.7 (0.8-37.4) | 28.2 (10.7-56.3) | 32.9 (20.0-49.1) | 42.8 (34.3-51.7) |
| Chandpur | 26.2 (7.9-59.5) | 25.8 (7.6-64.4) | 36.9 (17.0-62.6) | 43.5 (28.6-59.6) | 59.9 (40.2-76.8) |
| Chittagong | 9.6 (1.0-53.7) | 2.4 (0.2-20.1) | 15.4 (6.8-31.3) | 17.9 (7.8-36.1) | 48.2 (38.9-57.6) |
| Chuadanga | 0 | 51.7 (18.1-83.9) | 41.6 (22.8-63.2) | 83.6 (40.3-97.5) | 57.7 (7.8-95.7) |
| Cumilla | 17.0 (5.0-44.5) | 27.4 (11.1-53.5) | 38.6 (20.8-59.9) | 39.4 (23.9-57.4) | 52.5 (36.7-67.9) |
| Cox's Bazar | 5.6 (0.8-31.0) | 0 | 15.3 (4.5-41.1) | 14.0 (5.6-31.0) | 60.4 (46.2-73.1) |
| Dhaka | 0 | 52.8 (5.5-95.5) | 43.9 (20.8-70.0) | 24.8 (17.8-31.0) | 59.1 (51.4-66.4) |
| Dinajpur | 34.0 (15.7-58.7) | 41.9 (21.0-66.1) | 47.8 (26.9-69.5) | 35.5 (11.7-71.1) | 88.0 (44.4-98.5) |
| Faridpur | 22.3 (6.5-54.3) | 39.9 (15.3-70.9) | 30.5 (9.6-64.6) | 55.7 (33.8-75.6) | 69.1 (32.4-91.3) |
| Feni | 25.0 (25.0-25.0) | 17.7 (7.0-38.2) | 28.8 (24.9-33.1) | 30.3 (8.9-66.0) | 55.8 (31.9-77.3) |
| Gaibandha | 6.0 (0.9-30.9) | 18.1 (3.8-55.3) | 10.7 (2.1-39.7) | 36.3 (11.1-72.1) | 49.3 (26.8-72.1) |
| Gazipur | 0 | 33.1 (2.8-89.3) | 14.3 (1.5-65.2) | 17.7 (8.9-32.1) | 76.8 (60.6-87.7) |
| Gopalganj | 33.3 (33.3-33.3) | 17.9 (2.3-66.5) | 35.7 (3.2-90.3) | 66.7 (66.7-66.7) | 50.0 (50.0-50.0) |
| Habiganj | 5.5 (1.2-22.4) | 11.0 (6.0-19.2) | 30.2 (18.7-44.7) | 39.7 (14.9-71.3) | 35.8 (13.8-66.1) |
| Jamalpur | 20.6 (9.6-38.5) | 13.9 (5.8-29.8) | 21.7 (4.7-60.7) | 49.2 (25.7-73.1) | 60.3 (34.3-81.5) |
| Jessore | 48.5 (5.3-94.1) | 24.8 (8.9-52.6) | 47.4 (29.5-66.0) | 43.4 (18.2-72.6) | 81.9 (60.0-93.2) |
| Jhalokati | 11.3 (1.6-50.5) | 32.2 (15.0-56.1) | 55.2 (19.1-86.6) | 18.2 (4.5-51.0) | 60.0 (11.0-94.8) |
| Jhenaidah | 23.3 (4.8-64.8) | 56.7 (14.3-91.1) | 57.4 (32.3-79.2) | 32.6 (17.0-53.4) | 81.3 (26.9-98.1) |
| Joypurhat | 0 | 0 | 67.8 (44.7-84.6) | 0 | 85.0 (71.6-92.8) |
| Khagrachhari | 0 | 0 | 0 | 0 | 0 |
| Khulna | 0 | 17.1 (3.3-55.7) | 42.8 (17.6-72.4) | 45.8 (23.3-70.2) | 76.9 (65.5-85.4) |
| Kishoreganj | 16.7 (8.0-31.6) | 27.2 (9.2-58.1) | 30.7 (13.1-56.4) | 31.4 (24.8-38.9) | 76.9 (59.3-88.4) |
| Kurigram | 16.5 (8.5-29.6) | 19.0 (7.1-42.0) | 9.3 (1.4-43.6) | 49.2 (22.2-76.7) | 100 |
| Kushtia | 21.6 (12.7-34.3) | 52.8 (25.1-78.9) | 32.3 (11.5-63.7) | 55.7 (31.8-77.2) | 64.7 (22.5-92.1) |
| Lakshmipur | 0 | 6.5 (1.0-31.5) | 11.8 (7.6-17.8) | 59.7 (37.3-78.6) | 81.8 (26.4-98.3) |
| Lalmonirhat | 7.0 (1.7-24.8) | 0 | 58.4 (31.7-80.9) | 35.6 (12.6-67.9) | 60.4 (11.3-94.8) |
| Madaripur | 16.4 (6.2-38.2) | 0 | 49.2 (5.5-94.2) | 75.6 (55.6-88.4) | 73.5 (31.9-94.2) |
| Magura | 25.3 (6.8-61.4) | 21.8 (10.8-39.0) | 35.0 (16.1-60.2) | 0 | 100 |
| Manikganj | 0 | 20.7 (2.2-75.1) | 0 | 50.6 (19.9-80.8) | 0 |
| Maulvibazar | 5.2 (1.7-13.3) | 10.7 (2.4-36.5) | 8.9 (1.0-47.7) | 29.4 (14.9-49.7) | 56.6 (36.1-75.1) |
| Meherpur | 100 | 0 | 44.5 (18.3-74.2) | 79.0 (50.6-93.3) | 100 |
| Munshiganj | 0 | 0 | 42.1 (19.9-68.0) | 82.1 (59.7-93.4) | 62.5 (35.4-83.6) |
| Mymensingh | 16.7 (7.9-32.1) | 35.2 (24.0-48.2) | 37.5 (23.9-53.45) | 30.4 (19.1-44.6) | 44.6 (29.3-61.1) |
| Naogaon | 23.9 (4.2-69.4) | 37.1 (12.6-70.8) | 30.7 (9.8-64.5) | 74.4 (57.6-86.1) | 89.2 (48.8-98.6) |
| Narail | 20.8 (9.8-38.9) | 0 | 64.8 (33.5-87.0) | 49.0 (15.2-83.7) | 100 |
| Narayanganj | 0 | 0 | 0 | 37.1 (16.8-63.3) | 71.4 (48.1-87.0) |
| Narsingdi | 18.2 (1.3-79.3) | 0 | 15.0 (2.8-52.4) | 46.0 (22.1-71.9) | 90.1 (78.0-95.9) |
| Natore | 7.7 (0.8-46.8) | 67.8 (35.7-88.9) | 20.8 (4.9-57.2) | 50.2 (27.5-72.8) | 52.3 (29.1-74.5) |
| Chapai Nawabganj | 43.4 (10.9-82.7) | 27.2 (15.0-44.1) | 13.9 (6.2-28.2) | 10.1 (1.6-44.0) | 31.7 (3.8-84.6) |
| Netrakona | 9.9 (5.2-17.9) | 17.8 (8.0-34.8) | 29.2 (19.8-40.7) | 36.7 (14.7-66.2) | 100 |
| Nilphamari | 21.6 (8.6-45.5) | 3.5 (0.5-22.6) | 21.9 (4.5-62.6) | 36.4 (10.0-74.6) | 56.6 (9.2-94.4) |
| Noakhali | 0 | 18.2 (3.8-55.7) | 24.3 (10.5-46.6) | 17.6 (13.3-22.9) | 50.2 (33.9-66.4) |
| Pabna | 22.6 (9.1-46.1) | 17.0 (5.9-40.1) | 37.7 (16.6-64.7) | 24.2 (8.9-51.1) | 43.6 (16.1-75.7) |
| Panchagarh | 0 | 33.0 (15.8-56.5) | 0 | 0 | 100 |
| Patuakhali | 7.2 (2.3-20.1) | 21.6 (6.8-51.2) | 20.8 (6.8-48.6) | 27.9 (11.6-53.2) | 56.8 (32.6-78.1) |
| Pirojpur | 17.1 (7.0-36.0) | 27.2 (12.7-49.1) | 28.0 (17.5-41.7) | 55.7 (22.4-84.5) | 38.8 (27.3-51.6) |
| Rajbari | 0 | 39.4 (14.1-72.1) | 0 | 60.2 (48.7-70.7) | 82.7 (21.2-98.8) |
| Rajshahi | 41.6 (19.3-68.0) | 52.5 (29.6-74.4) | 63.9 (37.1-84.1) | 61.4 (37.0-81.2) | 76.4 (43.2-93.2) |
| Rangamati | 0 | 16.8 (3.7-51.4) | 0 | 50.0 (50.0-50.0) | 0 |
| Rangpur | 4.9 (0.6-32.6) | 21.2 (8.9-42.4) | 42.5 (24.8-62.4) | 63.4 (33.1-85.9) | 90.5 (55.6-98.6) |
| Satkhira | 12.9 (1.5-59.1) | 5.1 (0.8-27.2) | 69.5 (36.3-90.1) | 68.9 (26.1-93.3) | 59.8 (10.9-94.7) |
| Shariatpur | 0 | 13.1 (2.4-48.2) | 17.8 (6.6-40.0) | 50.5 (28.8-72.0) | 62.7 (8.7-96.8) |
| Sherpur | 0 | 11.9 (2.4-42.6) | 11.7 (1.9-47.9) | 43.8 (10.5-83.0) | 67.8 (31.6-90.5) |
| Sirajganj | 26.7 (6.2-66.6) | 25.3 (13.6-42.1) | 19.9 (9.0-38.4) | 55.7 (24.5-83.0) | 68.8 (15.7-96.3) |
| Sunamganj | 5.5 (1.6-17.2) | 16.8 (8.2-31.4) | 36.6 (22.9-52.9) | 28.1 (14.3-47.7) | 34.6 (16.4-58.8) |
| Sylhet | 6.9 (1.1-32.9) | 14.7 (7.6-26.4) | 19.9 (8.8-39.0) | 28.4 (17.9-41.9) | 64.8 (52.2-75.6) |
| Tangail | 16.3 (4.8-43.1) | 33.7 (18.9-52.6) | 41.0 (25.5-58.6) | 65.7 (42.0-83.6) | 50.0 (50.0-50.0) |
| Thakurgaon | 26.2 (15.1-41.3) | 64.4 (39.5-83.3) | 27.3 (5.8-69.6) | 34.9 (6.0-81.7) | 76.7 (37.7-94.7) |
| **Urban Bangladesh** | **10.4 (6.1-17.1)** | **27.3 (19.2-37.4)** | **33.8 (26.7-41.7)** | **33.6 (29.1-38.5)** | **61.7 (57.1-66.2)** |
| **Rural Bangladesh** | **13.4 (10.8-16.6)** | **21.9 (18.7-25.6)** | **30.6 (26.7-34.8)** | **41.0 (36.2-46.0)** | **61.0 (55.0-66.7)** |
| **Overall Bangladesh** | **13.1 (10.7-15.9)** | **22.4 (19.3-25.8)** | **31.1 (27.6-34.9)** | **38.3 (34.9-41.9)** | **61.5 (57.8-64.9)** |

### Predictors of cesarean section delivery in Bangladesh

Table 4 presents the factors associated with institutional delivery, population-level cesarean delivery, and institutional cesarean delivery that we examined by using multilevel mixed-effects binary logistic regressions. Aligned with the overall increase in CS, we found that, as compared to 1999/2000, the likelihoods of institutional delivery were 7.03 times higher (95% CI, 6.27-7.88), and institutional cesarean delivery was 5.32 times higher (95% CI, 4.36-6.48) in 2017/18. However, the likelihoods of population-level cesarean delivery showed an even higher increase during the same time period, with an 11.46 times higher likelihood (95% CI, 9.71-13.54). Similar patterns were identified in the predictors of institutional delivery, population-level cesarean, and institutional cesarean delivery. Women of comparatively higher age, higher educational attainment, and those not formally employed had higher likelihoods than their counterparts of institutional delivery and CS delivery—both at the population and institutional levels. In contrast, women with parity greater than 2, those with a female child as the index birth, and those living in rural areas had lower likelihoods of institutional delivery and cesarean delivery.

**Table 4:** Multilevel mixed-effects binary logistic regression model assessing the socio-demographic factors associated with cesarean delivery in Bangladesh, 1999/2000-2017/2018.

| **Characteristics** | Institutional delivery,<br>aOR (95% CI) | CS delivery at the population level,<br>aOR (95% CI) | CS delivery at the institutional level,<br>aOR (95% CI) |
| --- | --- | --- | --- |
| **Survey year** |  |  |  |
| 1999-2000 (ref) | 1.00 | 1.00 | 1.00 |
| 2004 | 1.03 (0.91-1.17) | 1.38 (1.14-1.68)*** | 1.59 (1.26-2.00)*** |
| 2007 | 1.42 (1.26-1.60)*** | 2.45 (2.05-2.93)*** | 2.57 (2.07-3.19)*** |
| 2011 | 3.02 (2.69-3.38)*** | 5.32 (4.48-6.31)*** | 3.82 (3.11-4.70)*** |
| 2014 | 4.33 (3.87-4.86)*** | 7.08 (5.98-8.38)*** | 4.05 (3.31-4.96)*** |
| 2017-2018 | 7.03 (6.27-7.88)*** | 11.46 (9.71-13.54)*** | 5.32 (4.36-6.48)*** |
| **Women's age at birth** |  |  |  |
| ≤19 years (ref) | 1.00 | 1.00 | 1.00 |
| 20-34 years | 1.48 (1.38-1.58)*** | 1.68 (1.54-1.82)*** | 1.51 (1.35-1.68)*** |
| ≥35 years | 2.12 (1.77-2.53)*** | 2.85 (2.29-3.55)*** | 2.14 (1.62-2.84)*** |
| **Women's education** |  |  |  |
| No education (ref) | 1.00 | 1.00 | 1.00 |
| Primary | 1.93 (1.73-2.16)*** | 2.00 (1.68-2.38)*** | 1.16 (0.93-1.45) |
| Secondary | 4.57 (4.11-5.09)*** | 5.08 (4.31-5.99)*** | 1.79 (1.46-2.20)*** |
| Higher | 17.81 (15.59-20.35)*** | 15.96 (13.37-19.05)*** | 3.00 (2.40-3.73)*** |
| **Women's formal employment status** |  |  |  |
| Employed (ref) | 1.00 | 1.00 | 1.00 |
| Unemployed | 1.47 (1.36-1.59)*** | 1.40 (1.28-1.54)*** | 1.18 (1.05-1.33)*** |
| **Parity** |  |  |  |
| 1-2 (ref) | 1.00 | 1.00 | 1.00 |
| >2 | 0.57 (0.53-0.61)*** | 0.50 (0.45-0.55)*** | 0.63 (0.56-0.71) |
| **Child's gender** |  |  |  |
| Male (ref) | 1.00 | 1.00 | 1.00 |
| Female | 0.87 (0.82-0.92)*** | 0.84 (0.78-0.91)*** | 0.89 (0.81-0.98)** |

| Place of residence |  |  |  |
| --- | --- | --- | --- |
| Urban (ref) | 1.00 | 1.00 | 1.00 |
| Rural | 0.41 (0.38-0.44)*** | 0.53 (0.49-0.57)*** | 0.95 (0.86-1.06) |
| **Note:** | ***p<0.01, **p<0.05, | aOR=adjusted odds ratio, | CI: Confidence intervals, Ref: Reference group. |

### Effects of cesarean section delivery on neonatal mortality in Bangladesh

The findings on the association between CS delivery and neonatal mortality are presented in Table 5. The crude association indicates a 27% decline in the likelihood of neonatal mortality among those delivered through CS. However, upon adjusting this association for the year of the survey and other socio-demographic factors, we found that this association became statistically insignificant. This lack of significance was also observed when we plotted CS rates and neonatal mortality rates across districts (Figure 2), revealing higher neonatal mortality rates in districts where CS rates were also elevated. However, despite this pattern, we identified a declining likelihood of neonatal mortality over the survey years with a lower likelihood (aOR, 0.70, 95% CI, 0.56-0.88) for the year 2017/18 compared to 1990/2000. Women of relatively older age, possessing higher education, not engaged in formal work, and those having a female child at the index births reported lower likelihoods of neonatal mortality. Conversely, women with a parity greater than two reported a higher likelihood of neonatal mortality.

**Figure 2:**
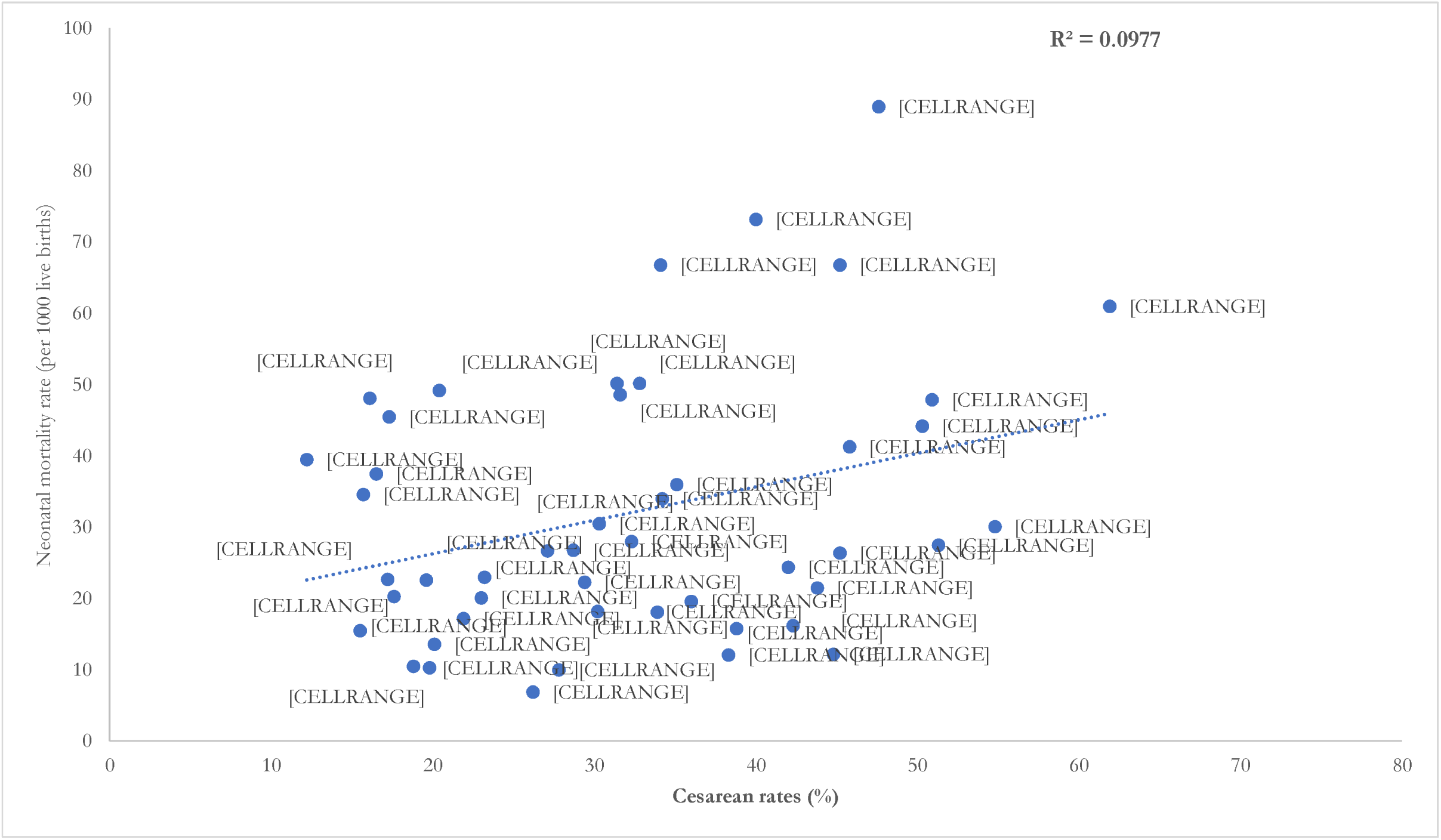
Cesarean section delivery rates (%) and neonatal mortality rates (per 1000 live births) in Bangladesh by districts (BDHS 2017/18)

**Table 5.**
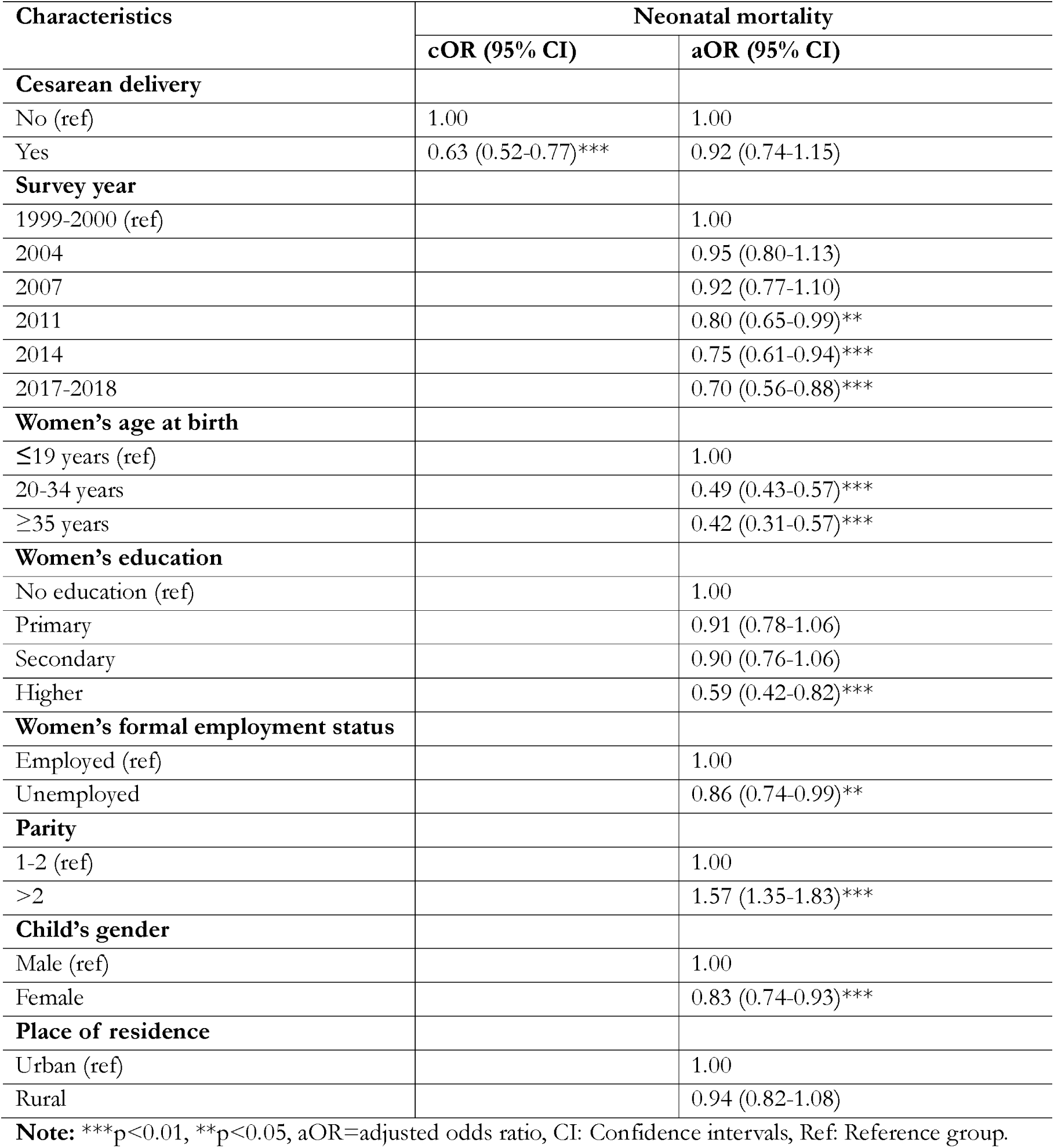
Results from multilevel mixed-effects binary logistic regression assessing the association between cesarean section delivery and neonatal mortality in Bangladesh, 1999/2000-2017/2018

| Characteristics | Neonatal mortality |  |
| --- | --- | --- |
|  | cOR (95% CI) | aOR (95% CI) |
| **Cesarean delivery** |  |  |
| No (ref) | 1.00 | 1.00 |
| Yes | 0.63 (0.52-0.77)*** | 0.92 (0.74-1.15) |
| **Survey year** |  |  |
| 1999-2000 (ref) |  | 1.00 |
| 2004 |  | 0.95 (0.80-1.13) |
| 2007 |  | 0.92 (0.77-1.10) |
| 2011 |  | 0.80 (0.65-0.99)** |
| 2014 |  | 0.75 (0.61-0.94)*** |
| 2017-2018 |  | 0.70 (0.56-0.88)*** |
| **Women's age at birth** |  |  |
| ≤19 years (ref) |  | 1.00 |
| 20-34 years |  | 0.49 (0.43-0.57)*** |
| ≥35 years |  | 0.42 (0.31-0.57)*** |
| **Women's education** |  |  |
| No education (ref) |  | 1.00 |
| Primary |  | 0.91 (0.78-1.06) |
| Secondary |  | 0.90 (0.76-1.06) |
| Higher |  | 0.59 (0.42-0.82)*** |
| **Women's formal employment status** |  |  |
| Employed (ref) |  | 1.00 |
| Unemployed |  | 0.86 (0.74-0.99)** |
| **Parity** |  |  |
| 1-2 (ref) |  | 1.00 |
| >2 |  | 1.57 (1.35-1.83)*** |
| **Child's gender** |  |  |
| Male (ref) |  | 1.00 |
| Female |  | 0.83 (0.74-0.93)*** |
| **Place of residence** |  |  |
| Urban (ref) |  | 1.00 |
| Rural |  | 0.94 (0.82-1.08) |

**Note:** \*\*\*p<0.01, \*\*p<0.05, aOR=adjusted odds ratio, CI: Confidence intervals, Ref: Reference group.

## Discussion

We explored a concerning trend in childbirth practices in Bangladesh. From 1999/2000 to 2017/18, there was a striking 42% increase in babies being born in hospitals, which is positive at first glance. However, when we looked closer, we found that CS deliveries increased by 30% during the same period. What is even more worrying is that the majority of this surge in hospital births was due to a substantial increase in CS performed in healthcare facilities. In 1999/2000, 30% of institutional deliveries involved CS, but by 2017/18, this had risen dramatically to 66%. Private healthcare facilities have emerged as major contributors to this rise in CS delivery, accounting for 80% of the country’s total CS delivery in 2017/18, compared to 45.5% in 1999/2000. Conversely, the rate of CS in government healthcare facilities dropped significantly from 49.7% in 1999/2000 to 15.5% in 2017/18. The situation becomes even more alarming when we examine district-level and wealth quintile-based variations. Women in border and hilly districts, as well as those in the poorest wealth quintile, reported significantly lower rates of CS delivery. In many cases, these rates fell below the 15% level. Conversely, in certain selected districts, and among women from affluent households, the reported usage of CS exceeded 80%. Our findings revealed no clear link between having a CS and neonatal mortality in Bangladesh, although the district-level Figure 2 suggests that neonatal mortality was higher in districts where CS rates were also higher. These findings raise concerns and warrant further research on the increasing use of CS and their effectiveness in improving child health outcomes in Bangladesh.

Since the 2000s, the Bangladesh government has prioritized increasing institutional deliveries and access to other maternal healthcare services [29]. This aligns with global goals like the Millennium Development Goals (MDGs) and Sustainable Development Goals (SDGs) and is common among LMICs [30]. However, despite significant efforts, progress has been limited. It is also important to acknowledge, ensuring universal coverage of institutional delivery, in line with the SDGs, is not practical for Bangladesh, as healthcare facilities cannot accommodate them all. As demonstrated by this and other studies in Bangladesh, half of all deliveries still occur outside formal healthcare facilities without skilled personnel present [31–34]. This study highlights another concerning trend: the rise in institutional deliveries is likely to be driven by an increase in CS use. While we cannot definitively say what proportion of these CS are unnecessary without further data, global evidence suggests a substantial proportion is likely to be provider-induced [35, 36]. This indicates that many women are choosing or being encouraged to undergo CS at healthcare facilities [29]. Whatever the reasons are, they are concerning. Women who choose CS mainly come from affluent wealth quintiles, urban areas, and are educated, or a combination of these factors [15]. This also indicates a pathway where the increasing demand for CS among this advantaged group is driving up its accessibility price, indicating that a segment of women cannot afford these services despite being in greater need due to the ongoing evidence of pregnancy complications in this group where CS is more warranted [15, 37]. The very high rate of CS among richer and urban women, along with a deficit of CS delivery among poorer women, supports this conclusion. Encouragement of CS delivery by healthcare facilities is linked with financial gains rather than its necessity [15]. This is the main reason for the rapid surge of CS in private healthcare facilities over the years, while public healthcare facilities observed a decline in CS, as seen in both this and other studies [15, 17, 29, 31, 33, 34]. However, whatever the direction, both indicate poor maternal and child health due to the unnecessary use of CS as well as an unmet need for CS [15, 17].

Concerns about the rising CS rate in Bangladesh surfaced in the early 2010s. National Safe Motherhood Guidelines and the Maternal Health Voucher Scheme were developed in response, followed by initiatives like enhanced skills training for healthcare professionals, public awareness campaigns, and financial incentives for promoting vaginal delivery [30, 38]. However, these programs yielded limited success in curbing the CS surge, coinciding with the increasing role of private healthcare facilities in providing CS [15]. Since 2020, the government has implemented a mandatory CS audit, requiring healthcare facilities to document the reasons for every CS performed [30]. This study reveals significant district-level variations in CS rates. The underlying reasons for these disparities are likely related to healthcare access, socioeconomic conditions, and cultural preferences [15, 17, 29]. Regions with better healthcare access and higher socioeconomic status tend to have higher CS rates, while rural and economically disadvantaged areas may experience lower rates due to accessibility and affordability challenges [15, 17]. Community awareness also play a role, with more informed populations potentially opting for CS based on health considerations [29]. Despite these diverse underlying factors, of the overall results suggest that universal policies and programs are a major drawback and indicate the need for more targeted approaches. Segmented policies tailored to specific local needs and contexts could be a more effective way to address the issue of unnecessary CS in Bangladesh. However, the country lags far behind in achieving this target.

While previous observations linked rising CS rates to lower neonatal mortality [39–41], Bangladesh’s current trend presents a puzzling contradiction. Despite a recent surge in CS deliveries, there is no clear association with reduced child mortality. Rather, there is an overall trend that districts with relatively high CS delivery rates tend to have elevated neonatal mortality. This suggests that the increased CS uptake may not be contributing to the intended goal of lowering child deaths. Potential explanations for this discrepancy could lie in two areas. First, advancements in the quality of maternal healthcare might be independently contributing to the decline in neonatal mortality, even as CS rates rise [42]. Second, the rise in CS use is mostly concentrated among women from affluent families, where neonatal mortality is generally low [15, 17, 29]. Since women from resource-poor families have higher neonatal deaths and they use CS much less than women from affluent households, the CS rate has a little or negative gradient [15–17, 29, 31]. These indicate a complex interplay among maternal healthcare services, CS rates, and neonatal mortality and warrant further research.

The findings of this paper have significant policy implications, particularly concerning the notable variations in CS rates among districts. It highlights deficits in CS use among women from resource-poor households in many districts, indicating that a one-size-fits-all approach to CS control may not be effective and could potentially worsen maternal and child health outcomes by limiting access for those in genuine need. The recommended approach is to shift focus towards promoting justified CS use, especially in private healthcare facilities where the majority of CS deliveries occur. This entails moving beyond the government’s objective of providing CS to enhance child health. The primary goal should be to ensure appropriate and justified CS utilization. In this endeavor, public healthcare facilities, despite their decreasing contribution to overall CS deliveries, can play a crucial role. By strengthening their capacity to provide high-quality, justified CS when needed, they can offer a safe and equitable alternative to private facilities, ultimately improving maternal and child health outcomes for all.

### Strengths and limitations

This paper’s major strength lies in the analysis of six rounds of nationally representative BDHS data, incorporating large samples. For the first time in Bangladesh, district-level data were generated for all waves of the BDHS survey, providing estimates for CS and relevant indicators. The analysis also presents CS variation across wealth quintiles, addressing deficits and excess use of CS. Advanced statistical modeling explored predictors of CS and the association between neonatal mortality and CS, considering sampling weights in all analyses. These comprehensive analyses provide substantial strength to leverage these findings for national-level policy and program development. However, the primary limitation is the use of cross-sectional data, indicating a correlational rather than causal nature of our findings. While geographic points were utilized to create district-level variables, a potential risk exists due to the displacement of the geographical location up to 5 km in rural areas and 2 km in urban areas, making the actual place of residence less evident. However, the BDHS approach ensured the displaced location remains within the original geographical location enhanced the validity of the district-level data. Since we had to extract district-level data by pooling geographical locations of survey data with district-level shapefiles, linking may not have been precise for the newly created districts. Moreover, many factors other than the ones available in the dataset that we considered may influence CS delivery, such as the distance of healthcare facilities.

## Conclusion

The rising trend in CS, particularly in private healthcare facilities, raises critical concerns in Bangladesh. The disproportionate distribution of CS across districts and socioeconomic strata highlights the need for a nuanced approach to childbirth practices. While government efforts to curb unnecessary CS have had limited success, the study suggests that a one-size-fits-all approach may exacerbate disparities. A shift in focus from merely increasing CS accessibility to ensuring justified and appropriate utilization and proactive role of public healthcare facilities in providing safe alternatives is recommended. Despite believing that the cesarean section is a lifesaving intervention for neonatal and maternal life, we also found that districts with an excessive cesarean rate also had higher neonatal mortality. All these indicate poor maternal and child health due to the unnecessary use of and unmet need for CS. Overall, the findings call for a reevaluation of existing regulations and policies to reduce the unnecessary use of CS. Targeted interventions, preferably at the district and lower tiers of local government levels, by appropriate regulatory bodies are needed to ensure transparent, need-based, and accountable cesarean deliveries to improve maternal and child health outcomes.

## Abbreviations

CS: Cesarean section
LMICs: Low- and Middle-Income Countries
WHO: World Health Organization
DHS: Demographic Health Survey
BDHS: Bangladesh Demographic Health Survey
NIPORT: National Institute of Population Research and Training
PSU: Primary Sampling Unit
aOR: adjusted Odds Ratio
cOR: crude Odds Ratio
CI: Confidence Interval
NGO: Non-Government Organizations
STROBE: Strengthening the Reporting of Observational Studies in Epidemiology.

## Declarations

### Author’s contribution

MNK developed the study concept. MBA and MAB performed the data analysis, MNK wrote the initial draft of the manuscript. SJK, MBA, MAB and MMI critically reviewed and edited the previous versions of the manuscript. All authors reviewed and approved the final version of the manuscript.

### Data availability

The datasets used and analysed in this study are freely available from the Measure DHS website: https://dhsprogram.com/. Interested persons are required to submit a research proposal to access the data for conducting this study, as we did.

### Ethics approval and consent to participate

The study data were sourced from the MEASURE DHS Archive, initially gathered by Macro in Calverton, USA. The data collection procedure received approval from the ORC Macro Institutional Review Board. Prior to enrolment, informed consent was obtained from all participants.

## Acknowledgement

We extend our gratitude to MEASURE DHS for their valuable data support. Additionally, the authors also acknowledge the support the Department of Population Science of Jatiya Kabi Kazi Nazrul Islam University, where the study was designed. We also acknowledge to the Health Systems and Population Studies Division of icddr,b, which acknowledges the support of the Governments of Bangladesh and Canada for providing core/unrestricted support for its operations and research.

## Supplementary files

**Supplementary table 1:**
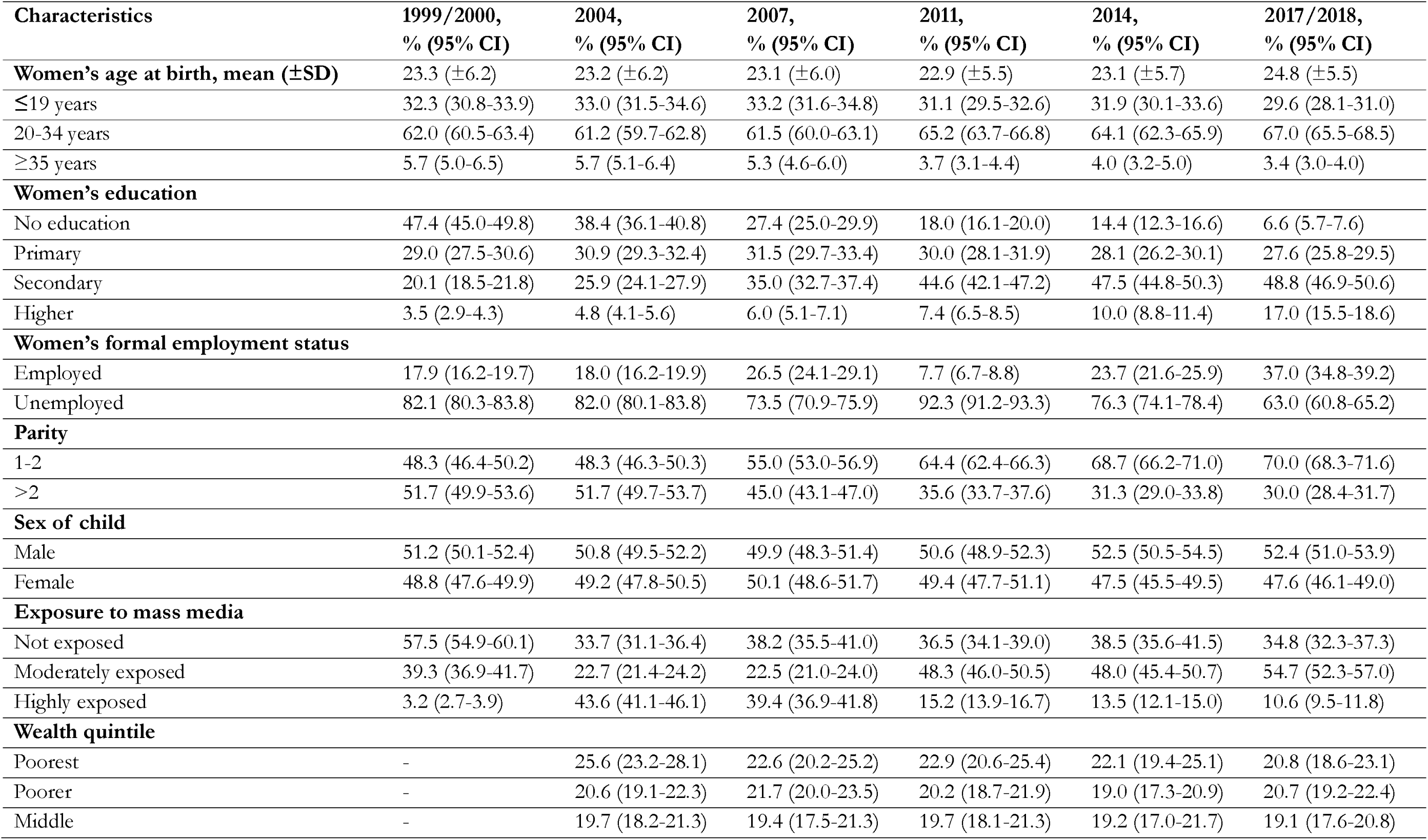
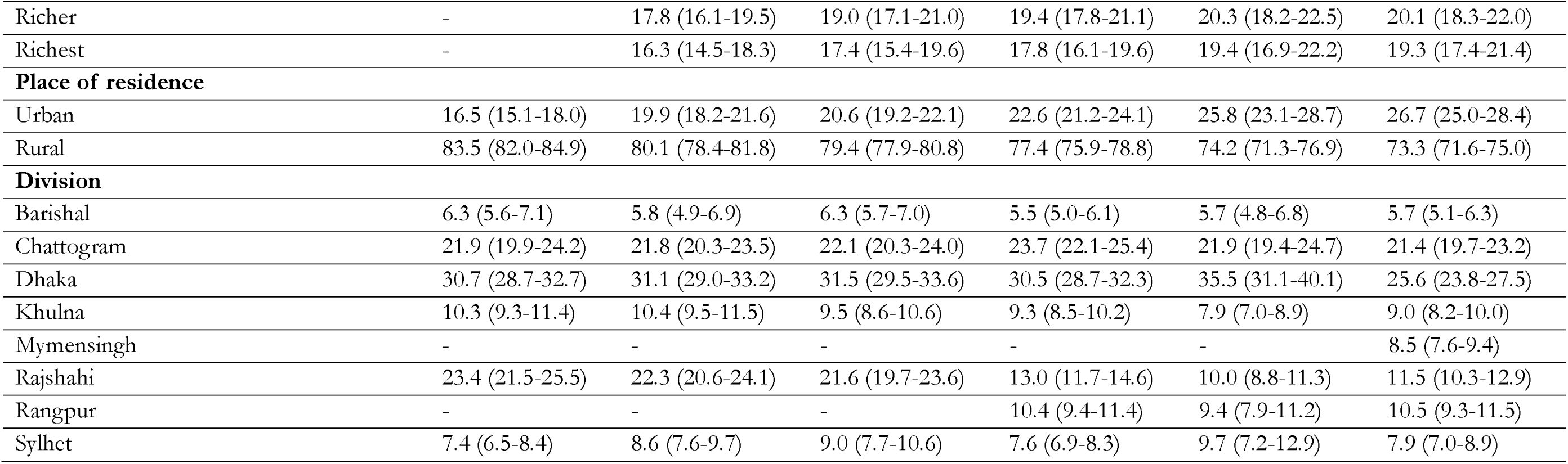
Background characteristics of the study respondents, 1999/2000-2017/18.

**Supplementary figure 1:**
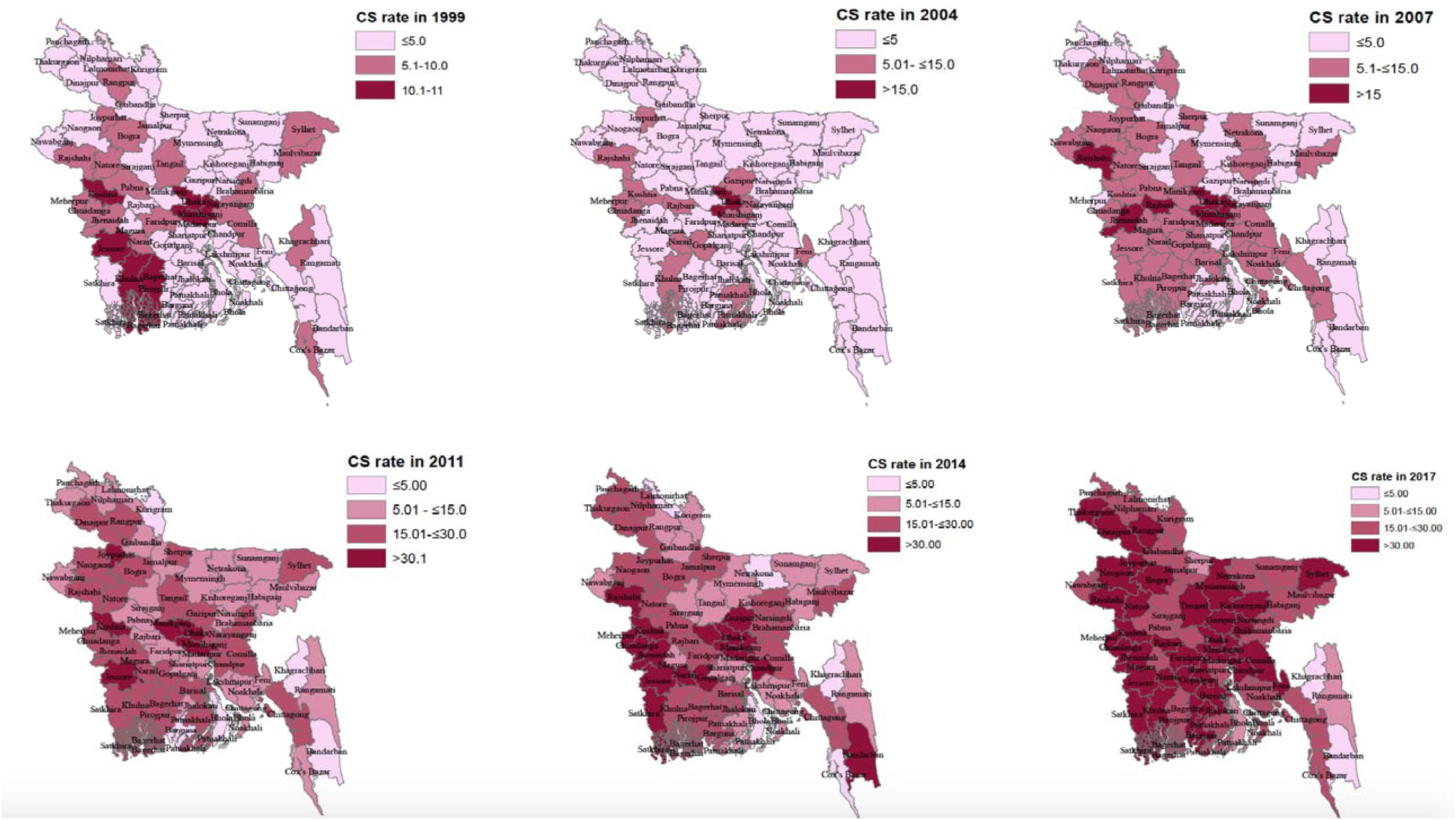
Trend in cesarean rate (CS) in Bangladesh from 1999/2000 to 2017/18.

**Supplementary figure 2:**
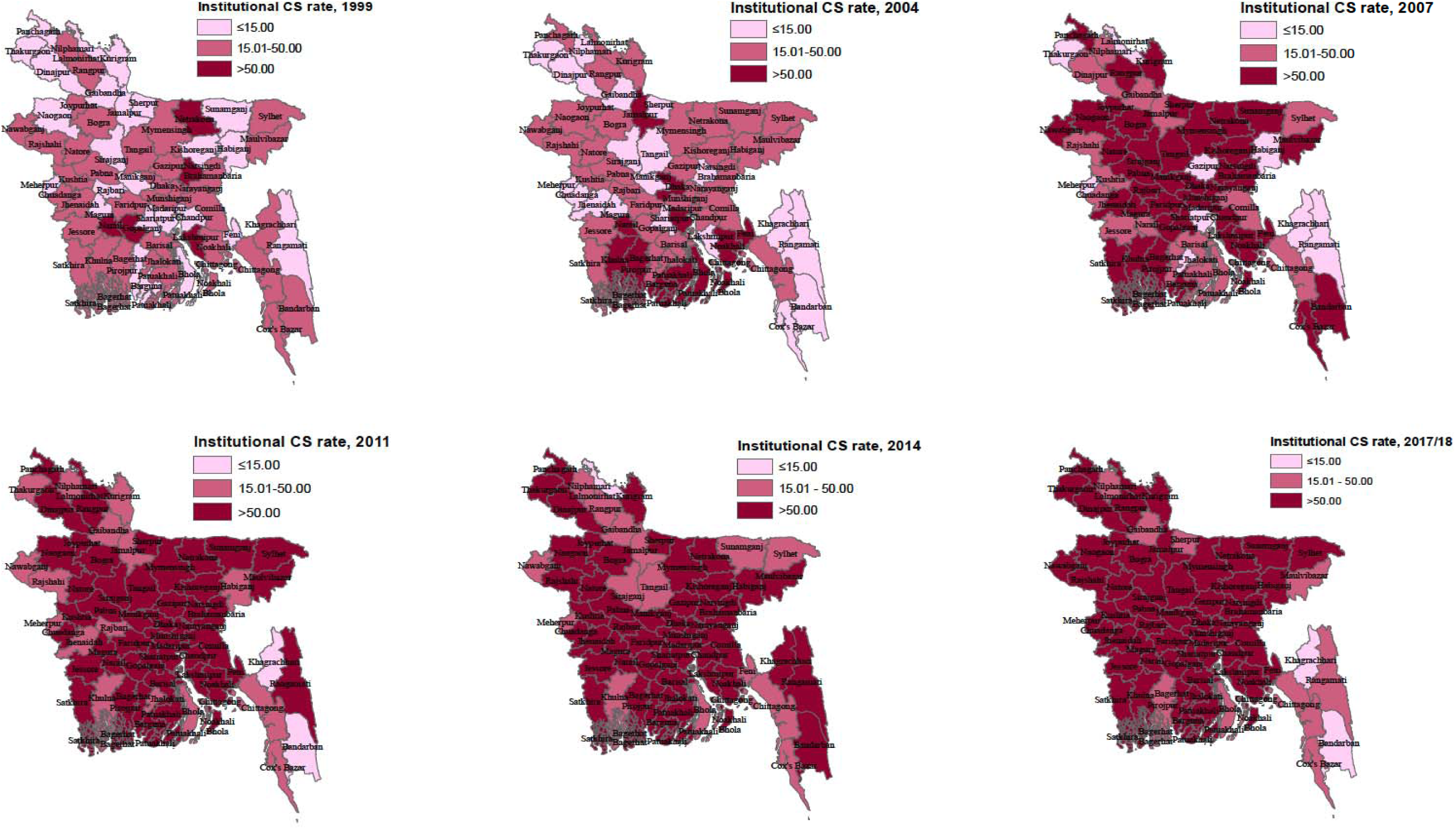
Trend in cesarean section (CS) rate in healthcare institutions in Bangladesh from 1999/2000-2017/2018.

**Supplementary figure 3:**
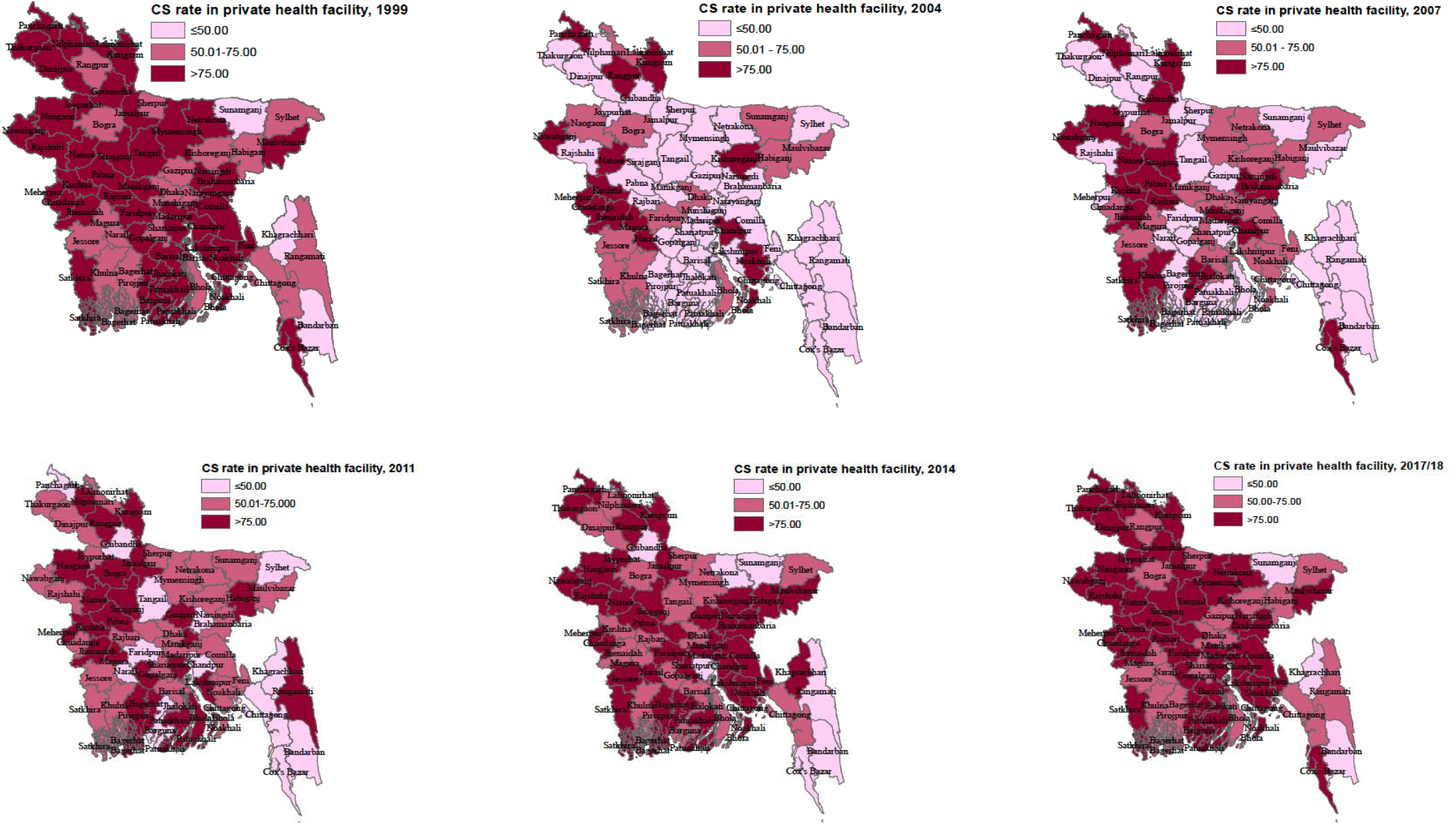
Trend in cesarean section (CS) rate in private healthcare facility in Bangladesh from 1999/2000-2017/2018.

**Supplementary table 2:** Population-based cesarean rates in Bangladesh by districts, Bangladesh, BDHS, 2014 and2017/18 (0-4% to 27.7%)

| **Districts** | **BDHS-2017/18, %** | **BDHS-2014, %** | **Difference between 2014 to 2017** |
| --- | --- | --- | --- |
| **Total** | **32.9** | **22.9** | **10.0** |
| Bagerhat | 23.0 | 22.4 | 0.6 |
| Bandarban | 0 | 50.0 |  |
| Barguna | 34.2 | 23.2 | 11.0 |
| Barishal | 36.0 | 29.9 | 6.1 |
| Bhola | 12.2 | 3.9 | 8.3 |
| Bogura | 30.3 | 20.3 | 10.0 |
| Brahmanbaria | 29.4 | 18.6 | 10.8 |
| Chandpur | 40.0 | 30.8 | 9.2 |
| Chittagong | 26.2 | 21.5 | 4.7 |
| Chuadanga | 54.0 | 43.9 | 10.1 |
| Cumilla | 38.8 | 28.1 | 10.7 |
| Cox's Bazar | 15.7 | 4.7 | 11.0 |
| Dhaka | 50.3 | 48.8 | 1.5 |
| Dinajpur | 44.8 | 22.6 | 22.2 |
| Faridpur | 39.2 | 24.6 | 14.6 |
| Feni | 34.1 | 6.2 | 27.9 |
| Gaibandha | 17.2 | 11.1 | 6.1 |
| Gazipur | 33.9 | 33.8 | 0.1 |
| Gopalganj | 36.6 | 39.8 | -3.2 |
| Habiganj | 15.5 | 8.1 | 7.4 |
| Jamalpur | 27.8 | 19.1 | 8.7 |
| Jessore | 51.3 | 36.8 | 14.5 |
| Jhalokati | 31.4 | 27.4 | 4.0 |
| Jhenaidah | 45.8 | 40.7 | 5.1 |
| Joypurhat | 47.6 | 23.4 | 24.2 |
| Khagrachhari | 0 | 4.5 |  |
| Khulna | 43.8 | 22.5 | 21.3 |
| Kishoreganj | 32.8 | 18.8 | 14.0 |
| Kurigram | 19.8 | 14.8 | 5.0 |
| Kushtia | 45.2 | 38.2 | 7.0 |
| Lakshmipur | 20.1 | 9.6 | 10.5 |
| Lalmonirhat | 18.8 | 2.5 | 16.3 |
| Madaripur | 41.7 | 11.1 | 30.6 |
| Magura | 28.7 | 13.7 | 15.0 |
| Manikganj | 26.9 | 30.4 | -3.5 |
| Maulvibazar | 17.3 | 22.8 | -5.5 |
| Meherpur | 67.5 | 34.2 | 33.3 |
| Munshiganj | 61.6 | 59.0 | 2.6 |
| Mymensingh | 32.3 | 13.2 | 19.1 |
| Naogaon | 50.9 | 26.4 | 24.5 |
| Narail | 36.2 | 32.2 | 4.0 |
| Narayanganj | 54.8 | 48.9 | 5.9 |
| Narsingdi | 42.3 | 20.6 | 21.7 |
| Natore | 41.8 | 29.4 | 12.4 |
| Chapai Nawabganj | 23.2 | 6.9 | 16.3 |
| Netrakona | 19.6 | 4.3 | 15.3 |
| Nilphamari | 22.8 | 19.9 | 2.9 |
| Noakhali | 21.9 | 14.8 | 7.1 |
| Pabna | 27.1 | 25.9 | 1.2 |
| Panchagarh | 16.1 | 30.1 | -14.0 |
| Patuakhali | 16.5 | 11.3 | 5.2 |
| Pirojpur | 30.2 | 19.1 | 11.1 |
| Rajbari | 61.9 | 21.5 | 40.4 |
| Rajshahi | 59.1 | 39.7 | 19.4 |
| Rangamati | 11.4 | 9.0 | 2.4 |
| Rangpur | 31.6 | 13.3 | 18.3 |
| Satkhira | 42.0 | 42.5 | -0.5 |
| Shariatpur | 20.4 | 12.6 | 7.8 |
| Sherpur | 10.2 | 24.6 | -14.4 |
| Sirajganj | 29.3 | 11.6 | 17.7 |
| Sunamganj | 17.6 | 6.8 | 10.8 |
| Sylhet | 35.1 | 12.7 | 22.4 |
| Tangail | 38.3 | 14.1 | 24.2 |
| Thakurgaon | 45.2 | 23.2 | 22.0 |

